# DNA methylation effects on Van der Woude Syndrome phenotypic variability

**DOI:** 10.1101/2023.11.04.23298094

**Authors:** Amanda Seaberg, Waheed Awotoye, Fang Qian, Lindsey Dunlay, Azeez Butali, Jeff Murray, Lina Moreno-Uribe, Aline Petrin

## Abstract

**OBJECTIVE:** Van der Woude Syndrome (VWS) classically presents with combinations of lip pits (LP) and orofacial clefts, with marked phenotypic discordance even amongst individuals carrying the same mutation. Such discordance suggests a possible role for epigenetic factors as phenotypic modifiers. Both *IRF6*, causal for 70% of VWS cases, and *TP63* interact in a regulatory loop to coordinate epithelial proliferation and differentiation for palatogenesis. We hypothesize that differential DNA methylation (DNAm) in CpG sites within regulatory regions of *IRF6* and *TP63* are associated with VWS phenotypic discordance.

**METHODS:** We measured DNAm levels of CpG sites located in the promoter regions of *IRF6* and *TP63* and in an *IRF6* enhancer element (MCS9.7) in 83 individuals with VWS grouped within 5 phenotypes for primary analysis: 1=CL+/-P+LP, 2=CL+/-P, 3=CP+LP, 4=CP, 5=LP and 2 phenotypes for secondary analysis: 1=any cleft and LP, 2= any cleft without LP. DNA samples were bisulfite converted and pyrosequenced with target-specific primers. Methylation levels were compared amongst phenotypes.

**RESULTS:** CpG sites in the *IRF6* promoter showed statistically significant differences in methylation among phenotypic groups in both analyses (P<0.05). Individuals with any form of cleft (Groups 1-4) had significantly higher methylation levels than individuals with lip pits only (Group 5). In the secondary analysis, individuals in Group 1 (cleft+LP) had significantly higher methylation than Group 2 (cleft only).

**CONCLUSION:** Results indicated that hypermethylation of the *IRF6* promoter is associated with more severe phenotypes (any cleft +/- lip pits); thus, possibly impacting an already genetically weakened *IRF6* protein and leading to a more severe phenotype.

## Introduction

Orofacial clefts are common birth defects affecting approximately 1 in 1,000 individuals, with prevalence varying by ethnicity and gender. While most forms of orofacial clefting are non-syndromic, approximately 30% are associated with other clinical signs and symptoms and are denominated syndromic cases ^1^. Van der Woude Syndrome (VWS), inherited through an autosomal dominant pattern, is among the most common syndromic forms of clefting, representing 2% of all orofacial clefts with a prevalence of 1 in 34,000 live births ^2^.

Mutations in the gene Interferon Regulatory Factor 6 (*IRF6*) are causal for 70% of VWS cases, while 5% of cases are caused by mutations in Grainyhead-Like Transcription Factor 3 (*GRHL3*) and the remaining 25% are of unknown cause. Causal mutations in *IRF6* commonly occur in the highly conserved DNA-binding domain (Exons 3 & 4) or the less conserved protein-binding domain (Exons 7-9) ^3,4^.

While these various established mutations in *IRF6* account for the presence of the syndrome, they do not fully explain the diverse phenotypic presentations of individuals with VWS. Lower lip pits (LP), occurring on the paramedian portion of the lower lip due to notching of the lips around day 36 of embryonic development, present in 85% of individuals with VWS ^5,6^. Other possible phenotypes include cleft lip with or without cleft palate (CL/P), cleft palate only (CPO) and hypodontia ^5^. Despite the autosomal dominant with high penetrance inheritance pattern, phenotype presentations for any given *IRF6*-causal mutation vary in multiple ways, even amongst family members^7^. This marked phenotypic discordance amongst individuals with VWS is exemplified by a 2011 study on a pair of monozygotic twins with variable phenotypic expression ^8^. One twin presented with bilateral cleft lip with cleft palate and bilateral lower lip pits, while the other twin presented with bilateral lower lip pits only. The occurrence of phenotypic discordance in these identical twins suggests a potential role for epigenetic factors as phenotypic modifiers in VWS.

One epigenetic factor that has been hypothesized as implicated in phenotypic discordance in VWS is DNA methylation (DNAm). A 2023 study by Petrin et al. reported findings of DNAm differences in individuals with VWS^4^. This paper reported methylation profiles of two pairs of monozygotic twins. The first pair was previously reported in the study that discovered *IRF6* as the first causal gene for VWS, while the second pair of twins is the pair reported in the aforementioned Jobling et al. paper where both individuals had VWS with discordant phenotypes ^4,7,8^. Whole genome DNAm was generated for both pairs of twins. While there were also other regions with differential methylation, Tumor Protein 63 (*TP63*) was the most significant hit amongst cleft related genes, showing DNAm differences between twin members. In the Kondo et al. twin set, the twin with VWS had higher methylation levels in several cpg sites located in or near *TP63* than the unaffected twin (delta beta from 5% to 9%); while in the Jobling et al. twin set, the twin with lip pits only had higher methylation levels in *TP63* than the twin with cleft lip and palate and lip pits (delta beta from 5% to 14%). They also observed one CpG site (cg26035071) in the promoter region of *IRF6* with higher absolute methylation levels in the twin with VWS in the Kondo et al. twin set (delta beta 9%) and in the twin with the more severe phenotype in the Jobling et al. twin pair^4^ (delta beta 10%).

Studies have shown a complex biological regulatory loop between *TP63* and *IRF6* in palatogenesis where normal levels of *IRF6* are necessary to downregulate *TP63*. However, the presence of an *IRF6* mutation can lead to aberrant DNAm of *TP63* which can modify phenotypic expression. In the Jobling twins, the higher methylation levels of *TP63* in the individual with lip pits only could have been enough to downregulate *TP63* during palatal fusion, allowing for effective palatal closure. Another possibility is that the higher methylation level in the promoter region of the *IRF6* gene, along with the causal mutation, contribute to a distinct phenotype. These hypotheses, along with the otherwise inexplicable phenotypic discordance observed in VWS, led to the development of this project.

In this article, we evaluated DNAm differences in three regions, *TP63* promoter, *IRF6* enhancer, and *IRF6* promoter, among individuals with *IRF6* causal mutations for VWS. We chose to analyze DNAm levels in *TP63* because of the DNAm differences found in the twins with VWS from Petrin et al. 2023. Also, we wanted to investigate if differential methylation in *IRF6* also leads to phenotypic heterogeneity. For *IRF6* we selected the *IRF6* immediate promoter and enhancer, MCS9.7. Of note a 2011 study showed that expression of *IRF6* is repressed by promoter methylation in individuals with squamous cell carcinoma ^9^. This finding points to a pathogenic effect led by silencing *IRF6* via DNA methylation of its promoter. The current study aims to explores the specific role of DNAm of *IRF6* and *TP63* on the phenotypic discordance observed in VWS.

## Methods

### Participants

We utilized blood or saliva DNA samples from 83 individuals with confirmed mutations in *IRF6* causal for VWS. For our primary analysis, these individuals were divided into five groups based on phenotype: Group 1 = CL/P and LP (n=46), Group 2 = CL/P (n=16), Group 3 = CPO and LP (n=10), Group 4 = CPO (n=6), Group 5 = LP (n=5). For our secondary analysis, the individuals were split into two groups: Group 1 = CL/P +LP or CPO + LP (n=56), Group 2 = CL/P only (n=22). All samples were obtained in accordance with prior study protocols, following their respective approval by their respective Institutional Review Boards (IRBs), and with informed consent provided by parents or guardians.

### Sample quality and bisulfite conversion

DNA quality was assessed and quantified with DropSense96™ and Qubit™ dsDNA High Sensitivity Range Assay Kit (ThermoFisher Scientific). After quantification of each specimen, 500 ng of genomic DNA were bisulfite converted using the EZ DNA Methylation™ Kit (Zymo Research, Irvine, CA, US) according to manufacturer’s protocol.

### Target Genes and DNA Amplification

We used the software PyroMark Assay Design 2.0 (Qiagen) software to design specific primers to amplify the CpG sites of interest in bisulfite converted DNA. Samples were amplified by PCR with designed primers for the identified target regions in the *IRF6* promoter, *IRF6* enhancer region, MCS9.7, and in the *TP63* promoter region. The *IRF6* promoter included ten target CpG sites, MCS9.7 included three target CpG sites, and the *TP63* promoter included two target CpG sites. Sample amplification was confirmed with agarose gel electrophoresis prior to pyrosequencing. Primer sequences and PCR conditions are available upon request.

### Pyrosequencing

Samples that passed the amplification quality check were then prepared for pyrosequencing. We used pyrosequencing primers, also designed with PyroMark Assay Design 2.0 (Qiagen) and specific to each target CpG site. The reactions were performed in a PyroMark Q48 Autoprep pyrosequencer and required reagents according to the manufacturer’s protocol. The resulting DNAm levels (%) of each CpG site for each sample was measured using the PyroMark Q48 AutoPrep software (Qiagen). Final methylation values (%) were exported to an excel spreadsheet and prepared for statistical analysis.

### Statistical Analysis

For the primary analysis, we conducted one-way ANOVA or one-way ANOVA on ranks as appropriate, followed by post-hoc Tukey-Kramer test, to assess the differences in DNAm levels among five phenotype groups. In the secondary analysis, we employed nonparametric Mann-Whitney U test to compare the DNAm levels between the two specific phenotype groups. Prior to performing the non-parametric statistical procedures, we conducted the Shapiro-Wilk test to evaluate the normality of the data distribution. A significance levels of 0.05 was applied to all statistical tests, and SAS® System v9.4, SAS Institute Inc. in Cary, NC, USA, was used for the statistical analysis.

## Results

### Primary Analysis

#### *IRF6* Immediate Promoter

Individuals in our cleft-affected groups, Groups 1, 2, 3 and 4, showed significantly higher methylation levels of the *IRF6* immediate promoter compared with individuals in Group 5. At CpG sites i6_6, i6_7, and i6_10, Groups 1 and 3 displayed significantly higher methylation levels than Group 5 (Table 2). However, no significant differences were found among phenotypes 1, 2, 3, and 4, or among phenotypes 2, 4, and 5 at these sites. At CpG site i6_8, Groups 1-4 all showed significantly higher methylation levels than Group 5. At site i6_9, Groups 1-3 displayed significantly higher methylation levels than Group 5, while no significant differences were found among phenotypes 1, 2, 3, and 4 or among phenotypes 4 and 5. Furthermore, there were no significant differences in methylation levels among Groups 1-4 at any CpG site.

**Table 1.** Distribution of sample sizes for primary and secondary analyses in subjects with Van der Woude Syndrome.

| Primary Analysis |  |  |  |
| --- | --- | --- | --- |
|  | *IRF6* Immediate Promoter | *IRF6* Enhancer, MCS9.7 | *TP63* |
| Group 1 (CL/P and LP) | 46 | 45 | 41 |
| Group 2 (CL/P) | 16 | 14 | 14 |
| Group 3 (CPO and LP) | 10 | 9 | 9 |
| Group 4 (CPO) | 6 | 5 | 5 |
| Group 5 (LP) | 5 | 5 | 5 |

| Secondary Analysis |  |  |  |
| --- | --- | --- | --- |
|  | *IRF6* Immediate Promoter | *IRF6* Enhancer, MCS9.7 | *TP63* |
| Group 1 (CL/P or CPO and LP) | 46 | 45 | 41 |
| Group 2 (CL/P or CPO only) | 16 | 14 | 14 |

**Table 2.** Comparison of *IRF6* value among phenotypes in subjects with Van der Woude syndrome.

| Variable/Phenotype | N | Mean±SD\Median | Minimum | Maximum | p-value |
| --- | --- | --- | --- | --- | --- |
| **i6_1:** |  |  |  |  | 0.081 |
| CL/P+LP | 46 | 0.39 ±0.24\0.33 | 0.05 | 1 |  |
| CL/P | 16 | 0.27±0.13\0.25 | 0.01 | 0.59 |  |
| CP+LP | 10 | 0.42±0.25\0.33 | 0.23 | 0.91 |  |
| CP | 6 | 0.40±0.29\0.27 | 0.16 | 0.82 |  |
| LP | 5 | 0.24±0.28\0.13 | 0.04 | 0.73 |  |
| **i6_2:** |  |  |  |  | 0.144 |
| CL/P+LP | 46 | 0.26±0.17\0.23 | 0.02 | 0.65 |  |
| CL/P | 16 | 0.19±0.09\0.18 | 0.05 | 0.42 |  |
| CP+LP | 10 | 0.27±0.16\0.21 | 0.13 | 0.58 |  |
| CP | 6 | 0.27±0.20\0.18 | 0.1 | 0.59 |  |
| LP | 5 | 0.16±0.22\0.07 | 0.03 | 0.54 |  |
| **i6_3:** |  |  |  |  | 0.202 |
| CL/P+LP | 46 | 0.40±0.25\0.35 | 0.06 | 1 |  |
| CL/P | 16 | 0.30±0.12\0.28 | 0.13 | 0.57 |  |
| CP+LP | 10 | 0.43±0.27\0.32 | 0.21 | 1 |  |
| CP | 6 | 0.39±0.27\0.28 | 0.17 | 0.87 |  |
| LP | 5 | 0.26±0.36\0.10 | 0.06 | 0.9 |  |
| **i6_4:** |  |  |  |  | 0.124 |
| CL/P+LP | 46 | 0.29±0.18\0.26 | 0.03 | 0.73 |  |
| CL/P | 16 | 0.21±0.09\0.19 | 0.07 | 0.43 |  |
| CP+LP | 10 | 0.31±0.19\0.25 | 0.16 | 0.69 |  |
| CP | 6 | 0.29±0.21\0.19 | 0.14 | 0.66 |  |
| LP | 5 | 0.17±0.22\0.07 | 0.04 | 0.57 |  |
| **i6_5:** |  |  |  |  | 0.113 |
| CL/P+LP | 46 | 0.26±0.17\0.22 | 0.03 | 0.71 |  |
| CL/P | 16 | 0.19±0.08\0.17 | 0.07 | 0.4 |  |
| CP+LP | 10 | 0.27±0.16\0.22 | 0.14 | 0.59 |  |
| CP | 6 | 0.26±0.20\0.17 | 0.1 | 0.61 |  |
| LP | 5 | 0.15±0.21\0.06 | 0.03 | 0.53 |  |
| **i6_6:** |  |  |  |  | 0.004 |
| CL/P+LP | 46 | 0.34±0.21\0.30* | 0.06 | 0.99 |  |
| CL/P | 16 | 0.26±0.12\0.24 | 0.13 | 0.58 |  |
| CP+LP | 10 | 0.37±0.22\0.30* | 0.19 | 0.81 |  |
| CP | 6 | 0.35±0.25\0.23 | 0.14 | 0.72 |  |
| LP | 5 | 0.08±0.06\0.09* | 0 | 0.15 |  |
| **i6_7:** |  |  |  |  | **0.004** |
| CL/P+LP | 46 | 0.14±0.09\0.12* | 0.01 | 0.37 |  |
| CL/P | 16 | 0.10±0.06\0.09 | 0.02 | 0.24 |  |
| CP+LP | 10 | 0.16±0.09\0.14* | 0.07 | 0.34 |  |
| CP | 6 | 0.16±0.14\0.08 | 0.05 | 0.36 |  |
| LP | 5 | 0.03±0.03\0.02* | 0 | 0.07 |  |
| **i6_8:** |  |  |  |  | **0.031** |
| CL/P+LP | 46 | 0.44±0.22\0.39* | 0.12 | 1 |  |
| CL/P | 16 | 0.36±0.14\0.37* | 0.22 | 0.76 |  |
| CP+LP | 10 | 0.45±0.24\0.35* | 0.26 | 0.95 |  |
| CP | 6 | 0.45±0.27\0.34* | 0.18 | 0.86 |  |
| LP | 5 | 0.16±0.12\0.15* | 0 | 0.3 |  |
| **i6_9:** |  |  |  |  | **0.010** |
| CL/P+LP | 46 | 0.37±0.21\0.31* | 0.08 | 0.86 |  |
| CL/P | 16 | 0.29±0.11\0.28* | 0.15 | 0.56 |  |
| CP+LP | 10 | 0.38±0.23\0.28* | 0.18 | 0.87 |  |
| CP | 6 | 0.36±0.20\0.27 | 0.17 | 0.66 |  |
| LP | 5 | 0.11±0.08\0.10* | 0 | 0.21 |  |
| **i6_10:** |  |  |  |  | **0.001** |
| CL/P+LP | 46 | 0.28±0.16\0.24* | 0.06 | 0.68 |  |
| CL/P | 16 | 0.19±0.07\0.18 | 0.11 | 0.34 |  |
| CP+LP | 10 | 0.29±0.15\0.22* | 0.17 | 0.59 |  |
| CP | 6 | 0.25±0.15\0.21 | 0.13 | 0.56 |  |
| LP | 5 | 0.09±0.06\0.09* | 0 | 0.17 |  |

\*For each variable, mean\median values with asterisks indicate significant differences ( $p < 0.05$ ) using Tukey-Kramer test.

#### *IRF6* Enhancer, MCS9.7

No significant differences in methylation levels were observed between any group at any of the three CpG sites, MCS_1, MCS_2, MCS_3, of the *IRF6* Enhancer, MCS9.7 (Table 3).

**Table 3.** Comparison of MCS9.7 values among phenotypes in subjects with Van der Woude syndrome

| Variable/Phenotype | N | Mean±SD\Median | Minimum | Maximum | p-value |
| --- | --- | --- | --- | --- | --- |
| **MCS_1:** |  |  |  |  | 0.523 |
| CL/P+LP | 41 | 0.04±0.03\0.02 | 0.01 | 0.11 |  |
| CL/P | 14 | 0.04±0.04\0.03 | 0.01 | 0.13 |  |
| CP+LP | 9 | 0.03±0.03\0.02 | 0.02 | 0.1 |  |
| CP | 5 | 0.05±0.03\0.04 | 0.02 | 0.09 |  |
| LP | 5 | 0.02±0.01\0.01 | 0 | 0.04 |  |
| **MCS_2:** |  |  |  |  | 0.143 |
| CL/P+LP | 41 | 0.86±0.29\1.00 | 0.12 | 1 |  |
| CL/P | 14 | 0.76±0.37\0.99 | 0.15 | 1 |  |
| CP+LP | 9 | 0.92±0.25\1.00 | 0.26 | 1 |  |
| CP | 5 | 1.00±0.00\1.00 | 1 | 1 |  |
| LP | 5 | 0.81±0.42\1.00 | 0.07 | 1 |  |
| **MCS_3:** |  |  |  |  | 0.284 |
| CL/P+LP | 41 | 0.76±0.28\0.93 | 0.1 | 1 |  |
| CL/P | 14 | 0.59±0.32\0.65 | 0.13 | 0.96 |  |
| CP+LP | 9 | 0.81±0.24\0.92 | 0.18 | 0.96 |  |
| CP | 5 | 0.84±0.11\0.85 | 0.67 | 0.95 |  |
| LP | 5 | 0.72±0.41\0.87 | 0 | 0.96 |  |

#### TP63

No significant differences in methylation levels were observed between any group at any of the two CpG sites, P63_1 and P63_2, of *TP63* (Table 4).

**Table 4.** Comparison of P63 gene mutation values among five phenotypes in subjects with Van der Woude syndrome.

| Variable/Phenotype | N | Mean±SD\Median | Minimum | Maximum | p-value |
| --- | --- | --- | --- | --- | --- |
| **P63_1:** |  |  |  |  | 0.643 |
| CL/P+LP | 45 | 0.66±0.25\0.72 | 0.11 | 1 |  |
| CL/P | 14 | 0.56±0.31\0.48 | 0.14 | 1 |  |
| CP+LP | 9 | 0.71±0.19\0.69 | 0.42 | 1 |  |
| CP | 5 | 0.59±0.25\0.57 | 0.3 | 1 |  |
| LP | 5 | 0.68±0.23\0.59 | 0.41 | 1 |  |
| **P63_2:** |  |  |  |  | 0.509 |
| CL/P+LP | 45 | 0.77±0.22\0.80 | 0.29 | 1 |  |
| CL/P | 14 | 0.69±0.21\0.65 | 0.4 | 1 |  |
| CP+LP | 9 | 0.82±0.16\0.89 | 0.59 | 1 |  |
| CP | 5 | 0.68±0.22\0.59 | 0.44 | 1 |  |
| LP | 5 | 0.69± 0.28\0.70 | 0.39 | 1 |  |

### Secondary Analysis

#### *IRF6* Immediate Promoter

Individuals in Group 1 displayed significantly higher methylation levels than individuals in Group 2 at CpG site 10 (p=0.011), as indicated in Table 5.

**Table 5.** Comparisons of the DNA methylation level between the two phenotypes among subjects with Van der Woude Syndrome.

|  | Phenotype I (n=54)<br>(CL+/-P or CP and LP) |  | Phenotype II (n=19)<br>(CL+/-P or CP only) |  | *P*-value |
| --- | --- | --- | --- | --- | --- |
|  | Mean ± SD\Median | Range | Mean ± SD\Median | Range |  |
| ***TP63*** |  |  |  |  |  |
| *TP63_1* | 0.67 ± 0.24\0.72 | 0.11-1.00 | 0.57 ± 0.29\0.54 | 0.14-1.00 | 0.184 |
| *TP63_2* | 0.78 ± 0.21\0.82 | 0.29-1.00 | 0.69 ± 0.21\0.65 | 0.40-1.00 | 0.086 |
| **MCS** | Phenotype I (n=50)<br>(CL+/-P or CP and LP) |  | Phenotype II (n=19)<br>(CL+/-P or CP only) |  | *P*-value |
|  | Mean ± SD\Median | Range | Mean ± SD\Median | Range |  |
| *MCS_1* | 0.04 ± 0.03\0.02 | 0.01-0.11 | 0.04 ± 0.04\0.03 | 0.01-0.13 | 0.757 |
| *MCS_2* | 0.87 ± 0.28\1.00 | 0.12-1.00 | 0.82 ± 0.33\1.00 | 0.15-1.00 | 0.174 |
| *MCS_3* | 0.77 ± 0.27\0.92 | 0.10-1.00 | 0.66 ± 0.29\0.79 | 0.13-0.96 | 0.056 |
| **i6** | Phenotype I (n=56)<br>(CL+/-P or CP and LP) |  | Phenotype II (n=22)<br>(CL+/-P or CP only) |  | *P*-value |
|  | Mean ± SD\Median | Range | Mean ± SD\Median | Range |  |
| *i6_1* | 0.40 ± 0.24\0.33 | 0.05-1.00 | 0.30 ± 0.19\0.25 | 0.01-0.82 | 0.060 |
| *i6_2* | 0.27 ± 0.17\0.22 | 0.02-0.65 | 0.21 ± 0.13\0.18 | 0.05-0.59 | 0.121 |
| *i6_3* | 0.41 ± 0.25\0.35 | 0.06-1.00 | 0.32 ± 0.17\0.28 | 0.13-0.87 | 0.167 |
| *i6_4* | 0.30 ± 0.18\0.26 | 0.03-0.73 | 0.23 ± 0.14\0.19 | 0.07-0.66 | 0.104 |
| *i6_5* | 0.27± 0.17\0.22 | 0.03-0.71 | 0.21 ± 0.13\0.17 | 0.07-0.61 | 0.086 |
| *i6_6* | 0.35 ± 0.21\0.30 | 0.06-0.99 | 0.28 ± 0.16\0.24 | 0.13-0.72 | 0.112 |
| *i6_7* | 0.15 ± 0.09\0.13 | 0.01-0.37 | 0.12 ± 0.08\0.09 | 0.02-0.36 | 0.073 |
| *i6_8* | 0.44 ± 0.22\0.39 | 0.12-1.00 | 0.39 ± 0.18\0.36 | 0.18-0.86 | 0.216 |
| i6_9 | 0.37 ± 0.21\0.31 | 0.08-0.87 | 0.31 ± 0.14\0.28 | 0.15-0.66 | 0.177 |
| i6_10 | 0.28 ± 0.15\0.24 | 0.06-0.68 | 0.21 ± 0.10\0.18 | 0.11-0.56 | **0.011\*** |

\* $p < 0.05$ considered statistically significant (obtained from the Wilcoxon sum-rank test)

#### *IRF6* Enhancer, MCS9.**7**

No significant differences in methylation levels were observed between the two groups at any of the three CpG sites in MCS9.7

#### TP63

No significant differences in methylation levels were observed between the two groups at either of the two CpG sites of *TP63*.

## Discussion

Despite mutations in *IRF6* representing a well-defined genetic cause for VWS, individuals with the syndrome display variable phenotypes. Epigenetic modifiers are thought to influence the variable phenotypic expression present in affected individuals.

We observed higher levels of DNAm in the *IRF6* immediate promoter with more severe phenotypes. Individuals in Groups 1-4 of the primary analysis all had some type of clefting, with or without lip pits, which resulted in higher methylation levels than individuals with lip pits only. Individuals with any type of clefting with lip pits also had higher methylation levels than individuals with any cleft only. This is thought to be due to the critical role that *IRF6* plays during lip and palate development, with normal levels of *IRF6* expression needed for proper elevation and fusion of the palatal shelves.

*IRF6* is part of a key biological regulatory loop with *TP63* which functions to help coordinate epithelial proliferation and differentiation during formation of the orofacial complex^9^. Disruption of this loop due to mutations in either gene can result in orofacial clefting. Expression of *IRF6* requires normal function of *TP63*, as *TP63* regulates *IRF6* expression by directly binding to two neighboring sites within MCS9.7: Motif1 and Motif2^10^. A study by Fakhouri et al. (2011) showed that abolishing both Motif1 and Motif2 significantly reduced and disrupted the MCS9.7 enhancer activity, whereas abolishing either motif separately reduced but did not disrupt enhancer activity ^11^.

Normal *IRF6* is expressed in high levels in the medial edge epithelia of the developing ectoderm, where it is an essential determinant of the keratinocyte proliferation-differentiation switch during lip and palate development^12-14^. High levels of *IRF6* also function indirectly to achieve palatal fusion by downregulating *TP63* at the time of palatal fusion. Downregulation of *TP63* at this point allows the pathway to enable disintegration of the median epithelial seam (MES) through apoptosis, allowing for mesenchymal continuity of the converging midfacial processes, resulting in successful palate closure ^10,15^. This negative feedback mechanism of *IRF6* on *TP63* was demonstrated in the mouse model, where *TP63* was spirited away at the time of palatal fusion in wild type mice while *IRF6* reached the pinnacle of its expression. Conversely, in mutant mice that were *IRF6* (-), the MES remained positive for *TP63* expression, resulting in a cleft and further indicating that the downregulation of *TP63* for palatal fusion necessitates normal *IRF6* function^10^. Mutations in *IRF6* causal for VWS can disrupt normal function of *IRF6*, hindering this important biological regulatory loop between *IRF6* and *TP63* and potentially resulting in orofacial clefting. *IRF6* Enhancer, MCS9.7, is the site where *TP63* binds and interacts with *IRF6*.

We hypothesize that, though the causal mutation acts to initially weaken the *IRF6* protein, methylation of the *IRF6* immediate promoter possibly compounds this by diminishing an already weakened protein, leading to a more severe phenotype, such as a cleft with lip pits. This follow-up study corroborates our previous findings that differential DNAm can act as modifiers influencing the severity of the phenotype.

Knowing that epigenetic changes can impact the process of craniofacial development, we now look at when these changes must occur. Epigenetic changes are impacted by several environmental factors, such as smoking, nutrition, stress, endocrine disruptors, etc. Given that the causal event for disruption of lip and palate formation occurs in-utero, these epigenetic changes likely occurred in either of the biological parents’ environments to alter the epigenetic makeup of their sperm and egg. Epigenetic mutations in the germline can be permanently programmed, potentially allowing for transgenerational transmission down the germline; however, extensive epigenetic reprogramming takes place upon fertilization^16,17^. This extensive reprogramming makes it difficult to determine whether these epigenetic mutations in the biological parents’ germline contribute to the methylation levels of *IRF6* observed in our study. Further studies are indicated to determine if factors in the parents’ environments contributing to epigenetic changes have a significant impact on craniofacial development.

There are some limitations in our study. To begin with, our groups were not evenly distributed in terms of the number of individuals, particularly in the case of Group 5, which exclusively consisted of individuals with lip pits. To better compare the methylation levels amongst phenotypes for VWS, we aim to increase our sample size in future studies. This will also be essential to determine if any true association exists between methylation of *TP63* or the *IRF6* enhancer, MCS9.7, and phenotype, as our study lacked the statistical power to draw this conclusion. Another limitation, common to many epigenetic studies, is that these samples were collected at a post-natal time point that does not correspond with the critical period of the disruption of lip or palate development, which occurred in-utero. Since epigenetic factors, such as DNAm, are influenced by environmental factors and can be altered throughout life, it is unclear whether the methylation levels observed in these individuals accurately corresponds with their methylation levels at the time of craniofacial development.

In the future, we aim to conduct a more extensive replication of this study with a larger sample size. This expanded research effort will enable us to robustly validate the assertions made in the original study and investigate the significance of MCS9.7 and *TP63* epigenetic influences. We chose to study individuals with *IRF6* mutations only, but we plan to extend the study to include individuals with *GRHL3* mutations in the future.

Genetic counseling and traditional genome sequencing provide a helpful, but incomplete picture for the basis of phenotypic inheritance and phenotypic divergence in Van der Woude Syndrome. This study lays the framework for connecting the *TP63-IRF6* signaling pathway and DNA methylation to the phenotypic divergence observed in VWS. Our study emphasizes the need for further understanding of the molecular mechanisms that regulate the epigenome and contribute to clinical presentation.

## Data Availability

All data produced in the present work are contained in the manuscript

## Acknowledgements

We would like to acknowledge Dr. Marie Gaine and Emese Kovacs of The University of Iowa Biomedical Science Program and Dr. Queena Lin of the Iowa NeuroBank Core for training us on use of the PyroMark Q48 Autoprep pyrosequencer.

